# Perspectives of persons with lived experience on acceptable outcome after traumatic brain injury

**DOI:** 10.1101/2025.02.21.25322625

**Authors:** Yelena G. Bodien, Lydia Borsi, Ellyn Pier, Samantha Kanny, Lillian Droscha, William Choi, Ryan Filoramo, Danielle Burnetta, Kathleen McColgan, Bhumi Patel, Mallory Spring, Jean Paul Vazquez Rivera, Jessica Wolfe, Enrico Quilico, Tiffany Campbell, Amanda R. Merner, Gabriel Lázaro-Muñoz, Lindsay Wilson, Joseph T. Giacino

## Abstract

**Importance:** Current approaches to functional outcome assessment in persons with severe traumatic brain injury (TBI) may not reflect the perspectives of persons with TBI or TBI caregivers.

**Objective:** Determine the lowest level of functional recovery after severe TBI that is perceived to be acceptable by persons with TBI and TBI caregivers.

**Design:** Cross-sectional crowdsourcing online survey disseminated May-July 2024.

**Setting:** United States.

**Participants:** Persons with a history of TBI requiring assistance with basic daily activities and TBI caregivers.

**Exposure(s):** History of severe TBI.

**Outcome(s)/Measure(s):** An expanded version of the Glasgow Outcome Scale-Extended (GOSE) designed to determine the level of acceptability of 11 TBI outcome milestones and the minimally acceptable outcome (MAO).

**Results:** The survey was completed by 252 persons with TBI (mean[SD] age 39.8[13.5] years; 67% female; 75% white; 11.9[12.0] years post-TBI) and 256 TBI caregivers (41.0[12.1] years; 57% female; 65% white). Among the outcomes selected most frequently as the MAO by persons with TBI (i.e., “recovery of yes/no communication” and “conscious, but does not communicate”) and TBI caregivers (i.e., “recovery of yes/no communication” and “alive, but permanently unconscious”), recovery of reliable yes/no communication was selected as acceptable by most respondents (persons with TBI: 36% vs 12%; Z=-7.1, p<0.0001; TBI caregivers 40% vs 14%, Z=-7.1, p=<0.0001). Recovery of reliable yes/no communication was therefore identified as the MAO by both cohorts. This outcome was rated as acceptable or somewhat acceptable by 65% of persons with TBI and 72% of caregivers. Outcomes representing disability greater than “completely independent in the home” were selected as the MAO more frequently than this common cut-off for “favorable” outcome, which was selected as the MAO by 5.6% and 3.9% persons with TBI and caregivers, respectively.

**Conclusions/Relevance:** Persons with TBI and TBI caregivers identified recovery of yes/no communication, an outcome that is well below the traditional cut-off for “favorable,” as the MAO. Persons with lived experience appear more accepting of a greater burden of disability than TBI investigators and providers. Recognizing this disparity in perspectives may influence clinical decision-making regarding goals of care and suggests the need for a more person-centered approach to TBI outcome assessment.

## INTRODUCTION

Severe traumatic brain injury (TBI) recovery trajectories vary widely.^1–3^ Among survivors, at least 20% remain with a severe disability.^3–5^ Medical decisions, including whether to prolong or withdraw life-sustaining treatment are anchored to the likelihood of attaining a “favorable” versus an “unfavorable” outcome.^6,7^ Although these terms permeate TBI literature and describe the endpoint for most clinical trials, their meaning is open to subjective interpretation and has not been vetted by the primary stakeholders – patients with TBI and TBI caregivers. These terms, which largely reflect the personal values of investigators and providers, may also have unintended adverse consequences on those directly impacted by TBI.

The Glasgow Outcome Scale-Extended (GOSE) is the most commonly-used global outcome measure in TBI research. This stems from the fact that the GOSE is the only TBI outcome measure designated as a “core” common data element by the National Institute of Neurological Disorders and Stroke, requiring its use across all TBI research studies.^8,9^ An 8-category ordinal scale, the GOSE is administered through a semi-structured interview that yields a total score ranging from 1 (death) to 8 (complete recovery). By design, most GOSE categories are broad. For example, the floor of the Lower Severe Disability category (GOSE 3) includes patients whose only sign of consciousness is visual pursuit while the ceiling is occupied by patients who can be left home alone safely for a few hours a day but require frequent assistance. While many TBI investigators have adopted the convention of dichotomizing the GOSE score into "favorable" or "unfavorable" outcome categories, there is no agreed upon cut-point between these two categories. A recent review found cut-points anywhere from a score of 2 to 7,^10^ although the upper boundary for “unfavorable” outcome is typically placed at a GOSE score of 3 or 4 (i.e., requires some assistance inside or outside the home).^2,3,9–11^ More importantly, it is unclear to what extent the outcomes represented in the GOSE are meaningful to persons with lived experience, as they were not formally engaged in the development process.

The aims of this study were to explore the perspectives of persons with TBI and TBI caregivers on the acceptability of graded TBI outcome milestones and to identify the minimally acceptable outcome (MAO). An “acceptable” outcome was defined as one that would be tolerable, even if it was not the one that would be most desired. We administered an on-line survey to both cohorts hypothesizing that the MAO selected by respondents would be “recovery to complete independence in the home” or worse.

## METHODS

### SPEAC-TBI Survey Development

We developed the Stakeholders’ Perspectives on Acceptable Outcomes after TBI (SPEAC-TBI, Supplementary Figure 1) survey which asks respondents to rate the acceptability of a graded series of TBI outcome milestones and to select the MAO. We used a three-step process to identify the 11 TBI outcomes. First, we expanded the eight existing GOSE categories to capture some additional incremental milestones. GOSE category 3 was expanded to include: 1) conscious but unable to communicate; 2) recovery of basic reliable yes-no communication; and 3) recovery of orientation. Second, we completed a scoping review to identify TBI outcomes that are not included in the GOSE. Six reviewers, in teams of two, reviewed 2,301 abstracts, conducted 162 full-text reviews, and extracted data from 123 qualifying studies. Conflicting ratings were reconciled by a third reviewer. No new outcomes were identified. Third, we invited persons with TBI, caregivers of persons with TBI, and TBI clinicians to participate in focus groups with the goal of ensuring that meaningful person-centered outcomes were included. Based on feedback acquired during the focus groups, we divided “independence in the home” into “partial” and “complete independence in the home.” We also added outcomes related to return to leisure activities, differentiating between leisure activities conducted alone and those conducted in a social setting. Focus group feedback also contributed to survey readability and refinement of the outcome descriptions. Table 1 describes the final set of 11 TBI outcome milestones used in the SPEAC-TBI survey.

**Table 1:**
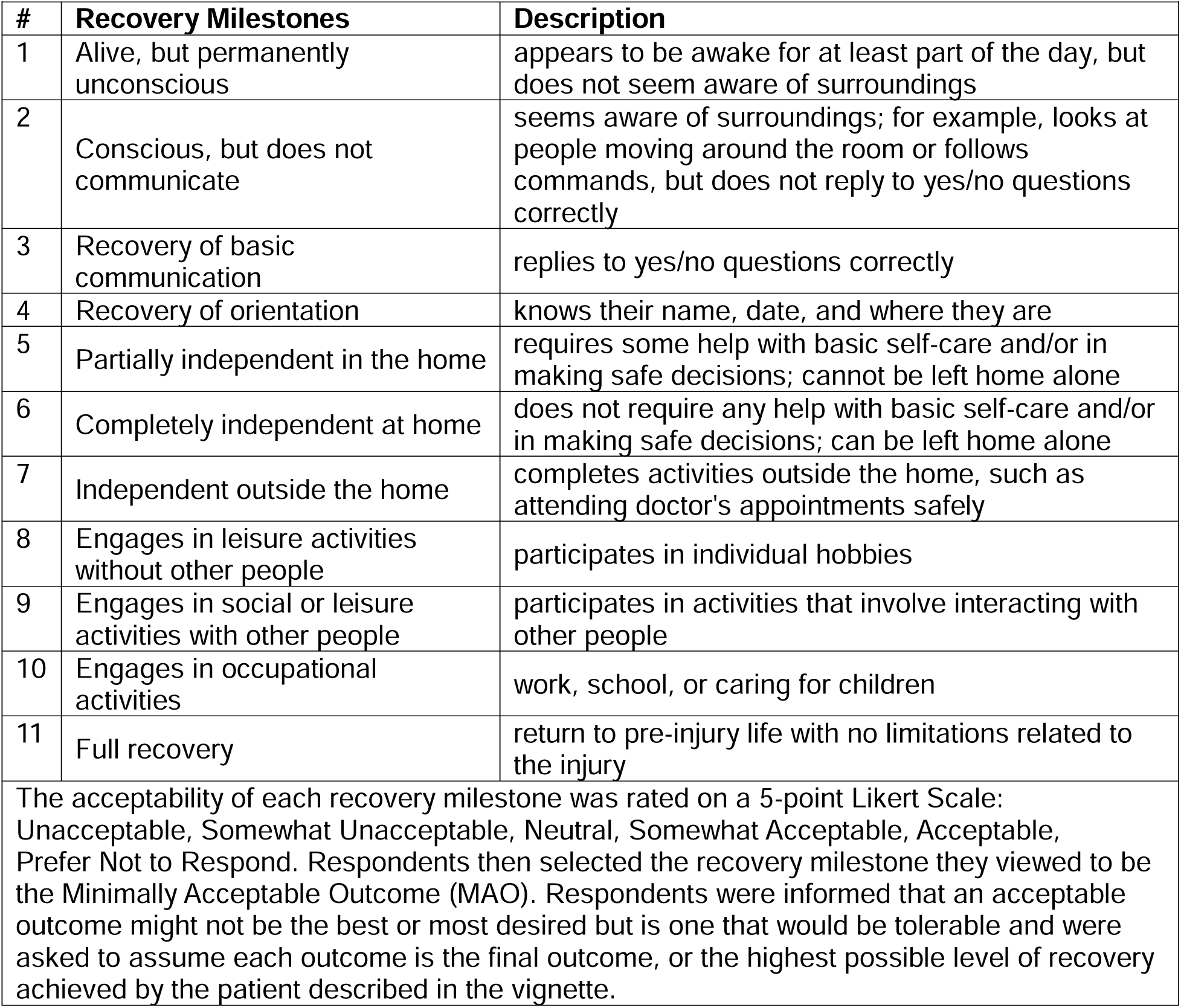
Recovery Milestones and Descriptions.

To anchor the survey questions, respondents were asked to read the following vignette and consider what an acceptable outcome would be for the person described in the vignette to assist with decision-making regarding goals of care:

> *Someone close to you had a serious car accident and got a severe traumatic brain injury. The person was unconscious when they arrived at the hospital. Their scans showed bruises and bleeding in the brain. The patient had emergency surgery to remove the blood in their brain. They are critically ill and have not improved in 3 days. They have not opened their eyes, responded to simple instructions, spoken, or moved on their own. It is now four days after the injury and the prognosis, or how the patient’s recovery will go, is very uncertain. You will need to make some decisions to help the doctors plan the patient’s care*.

Respondents were first asked to rate the 11 outcome milestones on a 5-point Likert-scale ranging from “unacceptable” to “acceptable” and then to select the MAO. A separate section of the survey asked respondents to rate the importance of personal values that might affect decision-making after a severe TBI (data not shown). The final SPEAC-TBI survey was created and administered in REDCap (Research Electronic Data Capture), a secure web-based application designed for creating and managing online surveys and research databases.^14,15^

### SPEAC-TBI Survey Distribution and Screening

The study was approved by the Massachusetts General Brigham Institutional Review Board. We distributed the REDCap SPEAC-TBI survey via Prolific, a crowdsourcing platform designed to rapidly yield a large sample of deidentified survey responses. Registered Prolific respondents are extensively vetted by Prolific personnel and their identities are verified, greatly minimizing the risk of fraudulent data. Respondents provided consent to participation after reviewing key information about the study and agreeing to complete the survey. We released the surveys in batches between May-July 2024 until our target enrollment of at least 250 participants in each cohort was met. We established this sample size based on the available number of Prolific respondents who endorsed TBI and an expected return rate of 50%, accounting for participants who did not meet inclusion criteria. Respondents were compensated at a rate of $12/hour in accord with Prolific’s recommendation.

We screened 215,332 Prolific participants to identify persons who endorsed a history of TBI on the following question provided by Prolific: *Do you have – or have you had – any condition, injury, or chronic illness?* Among Prolific users, 705 endorsed a history of TBI on this question. We also screened the 215,332 Prolific participants to identify persons who endorsed having caregiving responsibilities on the following question provided by Prolific: *I have extra responsibilities looking after a friend or member of my family due to a long-term physical or mental ill health or disability, or problem related to old age.* Among Prolific users, 12,862 endorsed being a caregiver on this question. Given that this question identifies caregivers but not specifically caregivers of persons with TBI, we distributed a follow-up screener survey to 4,098/12,862 (32%) caregivers to ask about the medical conditions of the person for whom they provided care. Of the 3,490 respondents, 321 (9.2%) endorsed caregiving for a person with TBI.

#### Inclusion Criteria– Persons with TBI

Persons with TBI, as identified by Prolific, were eligible for inclusion in SPEAC-TBI if they: 1) confirmed a history of TBI on the SPEAC-TBI survey and 2) endorsed requiring at least some assistance with daily activities due to the TBI. The latter criterion ensured that participants had a TBI-related disability, suggesting they sustained a severe TBI. We excluded participants who failed both of our attention check questions, which are used to verify response validity. We disseminated the SPEAC-TBI survey in batches and monitored responses until at least 250 respondents who met the inclusion criteria completed the survey. In total, 525 SPEAC-TBI surveys were disseminated to persons with TBI to achieve a final analytic sample of 252 respondents (Supplementary Figure 2A).

#### Inclusion Criteria– TBI Caregivers

TBI caregivers, as identified by Prolific and our additional screener question, were eligible for inclusion in SPEAC-TBI if they confirmed having a TBI caregiving role on the SPEAC-TBI survey. We excluded participants who failed both of our attention check questions. We disseminated the SPEAC-TBI survey to TBI caregivers in batches to 300 caregivers and monitored responses until at least 250 respondents who confirmed having a TBI caregiving role completed the survey. The final sample included 256 caregiver respondents (Supplementary Figure 2B).

### Statistical Analysis

We analyzed the TBI and caregiver respondent data separately. We used descriptive statistics to describe demographic characteristics. The MAO selected by persons with TBI and TBI caregivers was the primary outcome, and the acceptability ratings were the secondary outcomes. Our *a priori* analytic plan for identifying the MAO was based on a series of decisions described in Supplementary Figure 3. Briefly, we collapsed milestones that were selected as the MAO by fewer than 10 respondents, conducted a Chi-squared Goodness of Fit Test to determine whether there was a difference in the proportion of respondents selecting each outcome as the MAO, and performed a binomial pair-wise test between the two outcomes selected most frequently as MAOs. We then conducted a binomial pair-wise test to determine whether there was a difference in the number of respondents who selected “completely independent at home” as the MAO as compared to all other outcome milestones. We selected “completely independent at home,” as the reference because it is at the upper boundary of the GOSE “severe disability” category, and frequently the upper boundary of the dichotomized “unfavorable” outcome category.^10,16–18^ Finally, we compared the acceptability ratings of the outcomes selected most frequently as the MAO. We excluded the “Prefer not to respond” response option from our analyses. The statistical threshold was set at p<0.05. Analyses were conducted in SAS 9.4.

## RESULTS

### Participant Demographics

Persons with TBI (n=252) were mean (SD) age 39.8 (13.5) years; 67% (n=168) female; 75% (n=189) white; and 11.9 (12.0) years post-TBI. Half (n=125) were competitively employed, 12% (n=29) retired due to disability, and 47% (n=119) endorsed being religious (Table 2). Most respondents (n=199, 79%) reported that they needed help with daily activities due to the TBI “some of the time” and 21% (n=53) reported they needed help with daily activities “most of the time.” TBI caregivers (n=256) were mean (SD) age 41.0 (12.1) years; 57% (n=145) female and 65% (n=167) white. Most (n=180, 70%) were competitively employed and 51% (n=131) endorsed being religious (Table 2). Caregivers were predominantly children (n=87, 34%) or other first-degree relatives (n=77, 30%) of the TBI survivor.

**Table 2:** TBI and Caregiver Cohort Demographics.

| **Table 2: TBI and Caregiver Cohort Demographics** |  |  |
| --- | --- | --- |
| **Variable** | **TBI Cohort**<br>**N = 252** | **Caregiver Cohort**<br>**N = 256** |
| **Age; Mean (SD)** | 39.8 (± 13.5) | 41 (± 12.1) |
| **Sex, Female; N (%)** | 168 (66.7) | 145 (56.6) |
| **Race; N (%)** |  |  |
| American Indian/Alaska Native | 6 (2.4) | 3 (1.2) |
| Asian | 9 (3.6) | 13 (5.1) |
| Black or African American | 17 (6.7) | 59 (23.1) |
| White | 189 (75.0) | 167 (65.2) |
| Multiracial | 24 (9.5) | 10 (3.9) |
| Other | 4 (1.6) | 1 (0.4) |
| Prefer not to respond | 3 (1.2) | 3 (1.2) |
| **Marital Status; N (%)** |  |  |
| Single | 87 (34.5) | 74 (28.91) |
| Married/Living with Partner | 119 (47.2) | 157 (61.33) |
| Divorced/Separated | 33 (13.1) | 16 (6.3) |
| Widowed | 10 (4.0) | 6 (2.3) |
| Other | 1 (0.4) | 2 (0.8) |
| Prefer not to Respond | 2 (0.8) | 1 (0.4) |
| **Employment Status; N (%)** |  |  |
| Paid Employment | 125 (49.6) | 180 (70.3) |
| Retired (disability-related) | 29 (11.5) | 7 (2.7) |
| Retired (not disability-related) | 10 (4.0) | 3 (1.2) |
| Unemployed | 30 (11.9) | 11 (4.3) |
| Student (full-time and part-time) | 16 (6.3) | 21 (8.2) |
| Other | 37 (14.7) | 31 (12.1) |
| Prefer not to respond | 5 (2.0) | 3 (1.2) |
| **Educational Status; N (%)** |  |  |
| Less than high school | 3 (1.2) | 1 (0.4) |
| Completed high school | 34 (13.5) | 29 (11.3) |
| Working toward associate/vocational certificate | 23 (9.1) | 17 (6.6) |
| Completed associate/vocational certificate | 30 (11.9) | 30 (11.7) |
| Working toward college/university degree | 33 (13.1) | 32 (12.5) |
| Completed college/university degree | 73 (29.0) | 87 (34.0) |
| Working toward/completed masters/doctorate degree | 55 (21.8) | 58 (22.7) |
| Other | 1 (0.4) | 2 (0.8) |
| **Are you religious?; N (%)** |  |  |
| Yes | 119 (47.2) | 131 (51.2) |
| No | 123 (48.8) | 118 (46.1) |
| Prefer not to respond | 10 (4.0) | 7 (2.7) |

### Outcome Acceptability Ratings

All outcome milestones from “completely independent at home” up to and including “full recovery” were rated as acceptable by 80-85% of persons with TBI and 70-75% of TBI caregivers. However, 20% (n=50) of persons with TBI and 28% (n=57) of TBI caregivers reported that “alive and permanently unconscious” would be acceptable or somewhat acceptable (Figure 1, Supplementary Table 1). “Recovery of basic yes-no communication” was rated acceptable or somewhat acceptable by 65% (n=163) of persons with TBI and 72% of TBI caregivers (n=183). Outcome acceptability ratings were similar between persons with TBI who endorsed needing assistance with daily activities *all of the time* versus *some of the time* due to TBI (Supplementary Table 2).

**Figure 1:**
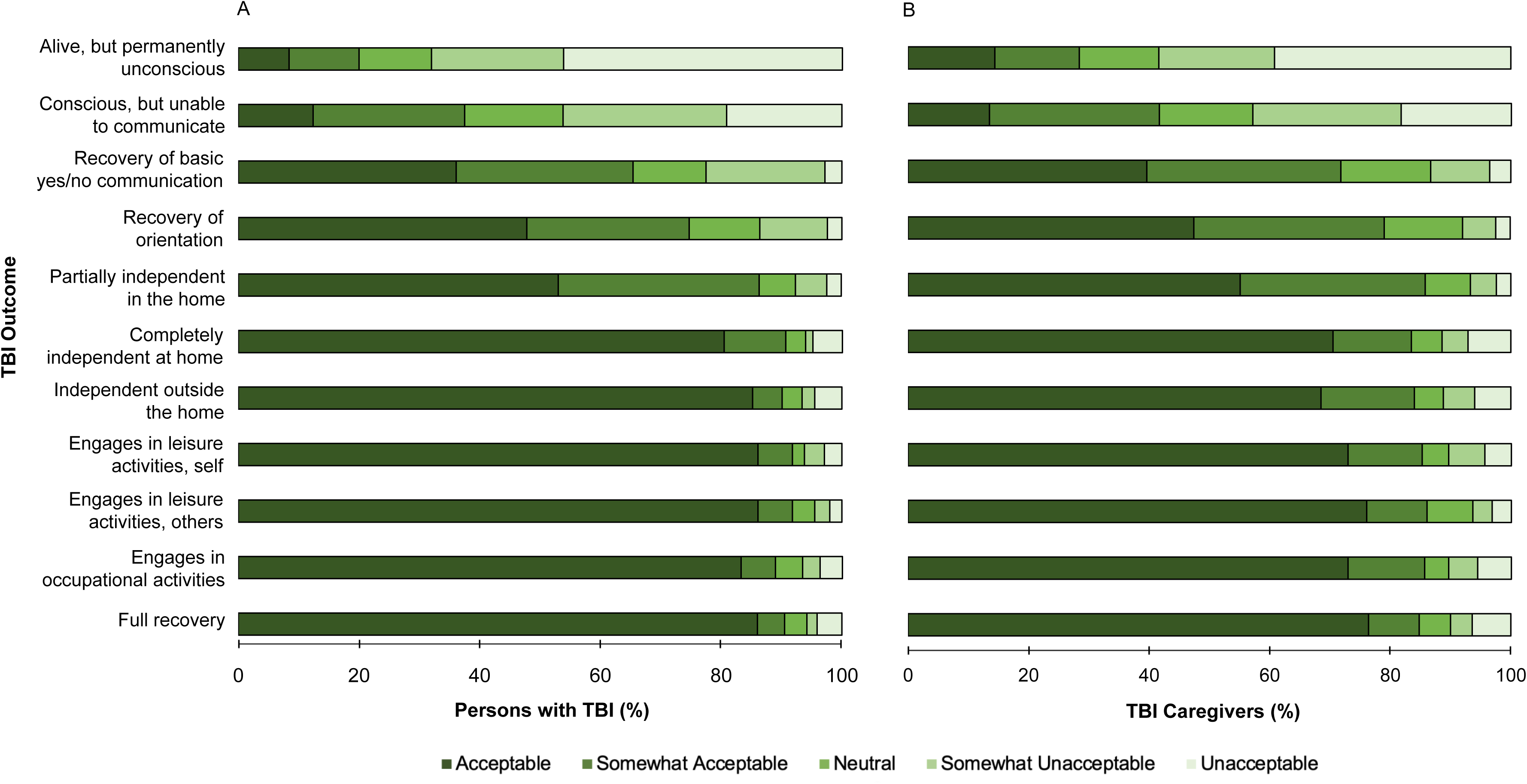
Outcome acceptability ratings. Shaded stacked bars represent acceptability ratings for persons with TBI (A) and TBI caregivers (B). Although the proportion of participants rating each outcome as acceptable generally increases as degree of disability decreases, a substantial proportion of persons with TBI (A) and TBI caregivers (B) rated lower-tiered outcomes associated with higher burden of disability as acceptable. Acceptability ratings were roughly equivalent for outcomes of recovery of independence outside the home through full recovery.

### Minimally Acceptable Outcome – Person with TBI

Fewer than 10 respondents selected “independent outside of the home,” “engages in leisure activities without other people,” “engages in social or leisure activities with other people,” “engages in occupational activities,” and “full recovery” as the MAO, therefore we collapsed these categories into a single category named “independent outside the home through full recovery” (Table 3, Supplementary Table 3). These results were similar between persons with TBI who endorsed needing assistance with daily activities *all of the time* versus *some of the time* due to TBI (Supplementary Table 4). The Chi-squared test comparing the frequency with which each recovery milestone was selected as the MAO was significant (X^2^=73.2, Df=6, p<0.0001, n=250; 2 responses of “prefer not to respond” were excluded) and all outcomes of greater disability than “completely independent at home” were selected as the MAO more often than this reference standard (Supplementary Table 5). However, there was no difference in the proportion of participants who selected the two most frequently selected MAOs, “conscious but does not communicate” (26%, n=65) and “recovery of basic yes-no communication” (22%, n=54; Z=1.0, p=0.31; Table 3, Figure 1).

**Table 3:**
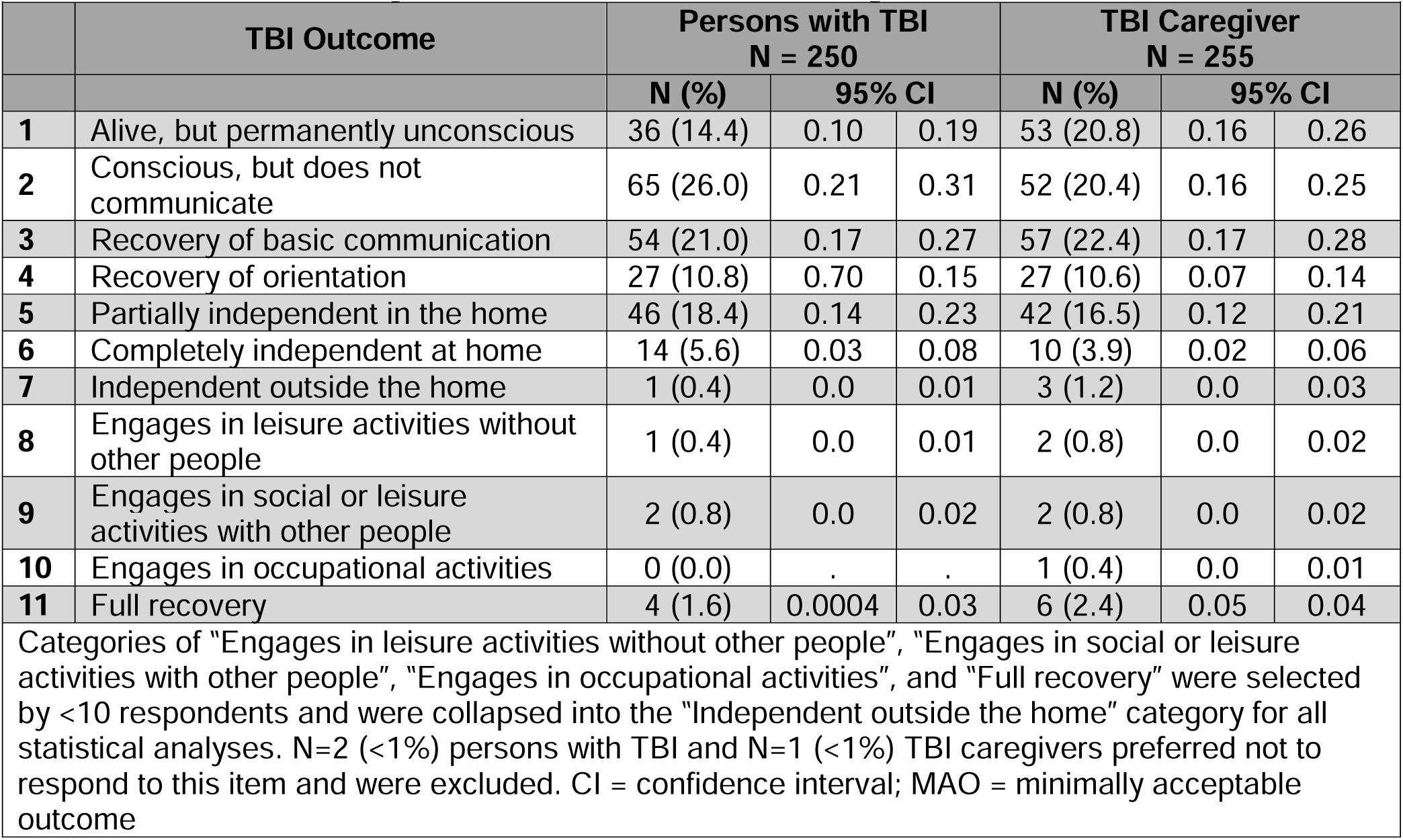
MAOs Selected by Persons with TBI and TBI Caregivers.

To differentiate “conscious but unable to communicate” and “recovery of basic yes-no communication” as the overall MAO (as endorsement rates were not significantly different), we relied on the standalone acceptability ratings of each of these outcomes. “Recovery of basic yes-no communication” was rated significantly more frequently as an acceptable outcome (36% vs 12%; Z=-7.1, p<0.0001) and significantly less frequently as an unacceptable outcome (3% vs 19%; Z=6.1, P<0.0001) than “conscious but unable to communicate” (Figure 1A, Supplementary Table 1). Based on these ratings, we established “recovery of basic communication” as the MAO for persons with TBI. This outcome was rated as acceptable or somewhat acceptable by 65% of persons with TBI (Supplementary Table 1).

### Minimally Acceptable Outcome – TBI Caregivers

Consistent with the data obtained from the persons with TBI cohort, fewer than 10 TBI Caregivers selected the five outcome categories from “independent outside the home” through “full recovery” as the MAO, therefore we collapsed these outcomes into a single category (Table 3, Supplementary Table 3). The Chi-squared test comparing the frequency with which each recovery milestone was selected as the MAO was significant (X^2^=62.1, Df=6, P< 0.0001, n=255; 1 response of “prefer not to respond” was excluded) and all outcomes of greater than disability than “completely independent at home” were selected as the MAO more often than this reference standard (Supplementary Table 5). However, there was no difference in the proportion of participants who selected each of the two most frequently selected MAOs, “recovery of basic yes-no communication” (22%, n=57) and “alive and unconscious” (21%, n=53; Z=-0.48, p=0.78; Figure 2, Table 3).

**Figure 2:**
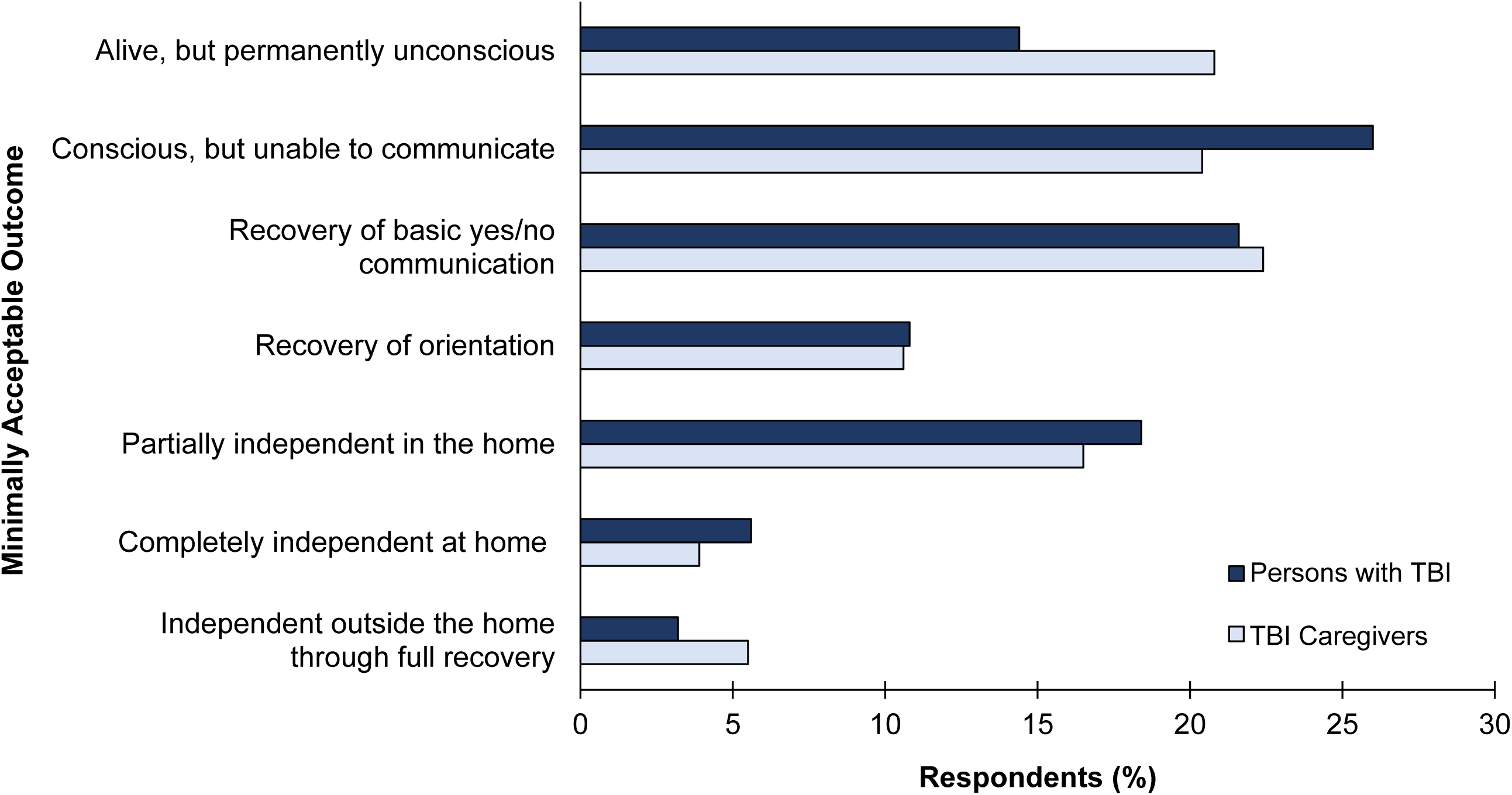
Minimally Acceptable Outcome (MAO) Ratings. Shading indicates the proportion of persons with TBI (dark blue) and TBI caregivers (light blue) who selected each outcome as the minimally acceptable outcome (MAO). Outcomes associated with lower disability burden (i.e., *“*independent outside the home” and better) were infrequently selected as the MAO and were collapsed into a single category named, *“*independent outside the home through full recovery.” Significantly more respondents from both participant groups selected outcomes associated with greater disability than “completely independent at home” (i.e., reference standard) as the MAO (Supplementary Table 5).

“Recovery of basic yes-no communication” was rated significantly more frequently as an acceptable outcome (40% vs 14%, Z=-7.1; p<0.0001) and significantly less frequently as an unacceptable outcome (4% vs. 39%, Z=9.2, p<0.0001) than “alive and unconscious” (Figure 1B, Supplementary Table 1). Based on these acceptability ratings, “recovery of basic communication” was established as the MAO for TBI Caregivers, matching the results in persons with TBI. This outcome was rated as acceptable or somewhat acceptable by 72% of TBI caregivers (Supplementary Table 1).

## DISCUSSION

In a survey of more than 500 respondents, persons with TBI and TBI caregivers selected “recovery of basic communication” as the minimally acceptable outcome (i.e., MAO) after severe TBI. When considering the acceptability of each outcome milestone separately, respondents selected outcomes associated with prolonged dependence (e.g., “alive, but not conscious”) as MAOs more frequently than those representing complete recovery of independence. These findings suggest that persons with lived experience value outcomes traditionally associated with high disability burden and view recovery that plateaus at the level of reliable communication as meaningful. Our results also suggest that prevailing approaches to how TBI outcome is conceptualized and measured should be person-centered.

Prior studies have found variability in the perspectives of TBI stakeholder groups on the acceptability of different levels of recovery after severe brain injury. Among surrogates of persons with acute neurological injury, recovery to GOSE 3 (i.e., requiring frequent assistance inside the home) warranted continuation of life-sustaining treatment. In contrast, healthcare professionals viewed GOSE 4 (i.e., requiring at least some assistance inside or outside the home) as the minimum level of recovery required to justify continuation of life-sustaining treatment.^19^ In separate studies, 65% of TBI caregivers rated a GOSE score of 3 as being worse than death,^20^ and more than 60% of TBI clinicians reported that a final outcome of GOSE 3 justifies withdrawing life-sustaining treatment.^21^ The GOSE 3 category is broad, encompassing persons who are minimally responsive and those who can be left home alone safely for a few hours per day.^22,23^ Disparate findings related to the acceptability of a GOSE 3 outcome may reflect variance in how this level of function is described in each study. In a study that did not rely on the GOSE, 82% of families of patients with acute severe brain injury identified “able to think and communicate" as a minimally acceptable recovery.^24^ This result is consistent with our observation that outcomes historically considered to be unfavorable may be personally meaningful to persons with lived experience.

Few existing outcome measures include item content that specifically addresses recovery of reliable communication. The Coma Recovery Scale-Revised probes communication ability, but is limited to yes-no situational orientation questions,^25^ which may not reflect the preferences of persons with TBI and their caregivers. The Disability Rating Scale evaluates expressive language function, ranging from no verbal or gestural signs of communication to accurate verbal expression of orientation, but does not assess the ability to reliably answer yes-no questions, which frequently recovers ahead of orientation. Indeed, none of the Curing Coma Campaign’s recently recommended Disorders of Consciousness Common Data Elements for Outcome and Endpoints capture recovery of basic communication,^26^ although tools under current development may soon fill this gap. The findings from this study can guide the development of new outcome measures that have strong construct validity and align with the preferences of persons with lived experience.

### Limitations

There are some limitations of this study that should be considered. First, an electronic survey format precludes confirmation of TBI injury severity, degree of caregiving, and other factors that characterize the sample. To ensure that responders had a significant TBI with injury characteristics that could have been similar to the survey vignette, we selected participants who endorsed needing at least some assistance with daily activities due to the TBI. However, we were unable to confirm other aspects of the injury severity. Our survey could only be completed by persons with TBI who had the ability to respond to questions, which excludes individuals with the most severe injuries who may hold different perspectives. The caregiver respondents likely represented persons with a wider range of TBI severity. Second, participants were predominantly female, white, and middle-aged, which is not representative of those at highest risk for TBI – namely, males, older individuals, and racial/ethnic minorities.^27^ Prolific survey respondents are likely to be more proficient with technology and, therefore, younger. The demographic composition of our caregiver sample is generally consistent with prior studies.^28^ Third, we were unable to probe participants’ rationale for selecting specific recovery milestones as MAOs. This requires a qualitative study design, which was not possible using the crowdsourcing platform. Finally, we anchored the survey to a single vignette, which may have elicited responses specific to the circumstances described in that particular scenario.

## CONCLUSION

In this study exploring the perspectives of persons with lived experience on the acceptability of tiered outcome milestones reflecting different degrees of recovery after severe TBI, we found that both persons with TBI and TBI caregivers viewed outcomes traditionally associated with severe disability as acceptable. Moreover, “recovery of basic communication” was selected as the minimally acceptable outcome (MAO), which is well below “complete recovery of independence,” the traditional cut-off for “favorable” outcome used in most TBI outcome studies. These findings suggest that persons with lived experience are more accepting of a greater burden of disability than TBI investigators and providers. It is important to recognize this disparity in perspectives as it may complicate clinical decision-making with regard to establishing goals of care and suggests the need for a more person-centered approach to TBI outcome assessment.

## Data Availability

All data produced in the present study are available upon reasonable request to the authors

## Acknowledgement

Author contributions: Conception or design (YGB, LB, JTG), data acquisition or analysis, (YGB, LB, EP, SK, LD, RF), data interpretation (YGB, LB, EP, SK, LD, DB, KM, BP, MS, JPV, JW, EQ, GLM, ARM, TF, LW, JTG). All authors contributed to drafting or reviewing the manuscript critically for important intellectual content and approved the final version to be published. YGB, LB, EP, RF, SK, LD, and JTG had access to the data for the duration of the study.

This study was funded by National Institute on Disability, Independent Living, and Rehabilitation Research grant 90DPTB0027 to Spaulding-Harvard TBI Model System.

The funders had no role in the design and conduct of the study; collection, management, analysis, and interpretation of the data; preparation, review, or approval of the manuscript; and decision to submit the manuscript for publication.

Disclaimer: The views expressed herein are those of the authors and do not reflect the official policy or position of the NIDILRR or HHS and do not constitute an endorsement by NIDILRR, HHS, or other components of the US federal government

## Notes

### Competing Interest Statement

The authors have declared no competing interest.

### Funding Statement

This study was funded by NIDLIRR

### Author Declarations

The Mass General Brigham gave ethical approval for this work

