## Supplementary Materials for "Perspectives of persons with lived experience on acceptable outcome after traumatic brain injury"

| **Table of Contents** | **Page** |
| --- | --- |
| Supplementary Figure 1: Survey Excerpts | 2 |
| Supplementary Figure 2: Analytic Decision Tree | 3 |
| Supplementary Figure 3: Analytic Decision Tree | 4 |
| Supplementary Table 1: Acceptability Ratings for Respondents with TBI and Caregivers | 5 |
| Supplementary Table 2: Acceptability of Outcomes Rated by Respondents with TBI Endorsing Needing Help with Basic Activities Some of the Time Versus All of the Time | 6 |
| Supplementary Table 3: MAOs selected by Respondents with TBI and Caregivers With Low Response Categories Collapsed | 8 |
| Supplementary Table 4: MAOs Selected by Respondents with TBI Endorsing Needing Help with Basic Activities Some of the Time Versus All of the Time | 9 |
| Supplementary Table 5: Result of Binomial Tests Comparing the Proportion of Participants Selecting Each TBI Outcome as the MAO to the Reference Outcome of “Completely Independent at Home” | 10 |

**Supplementary Figure 1**
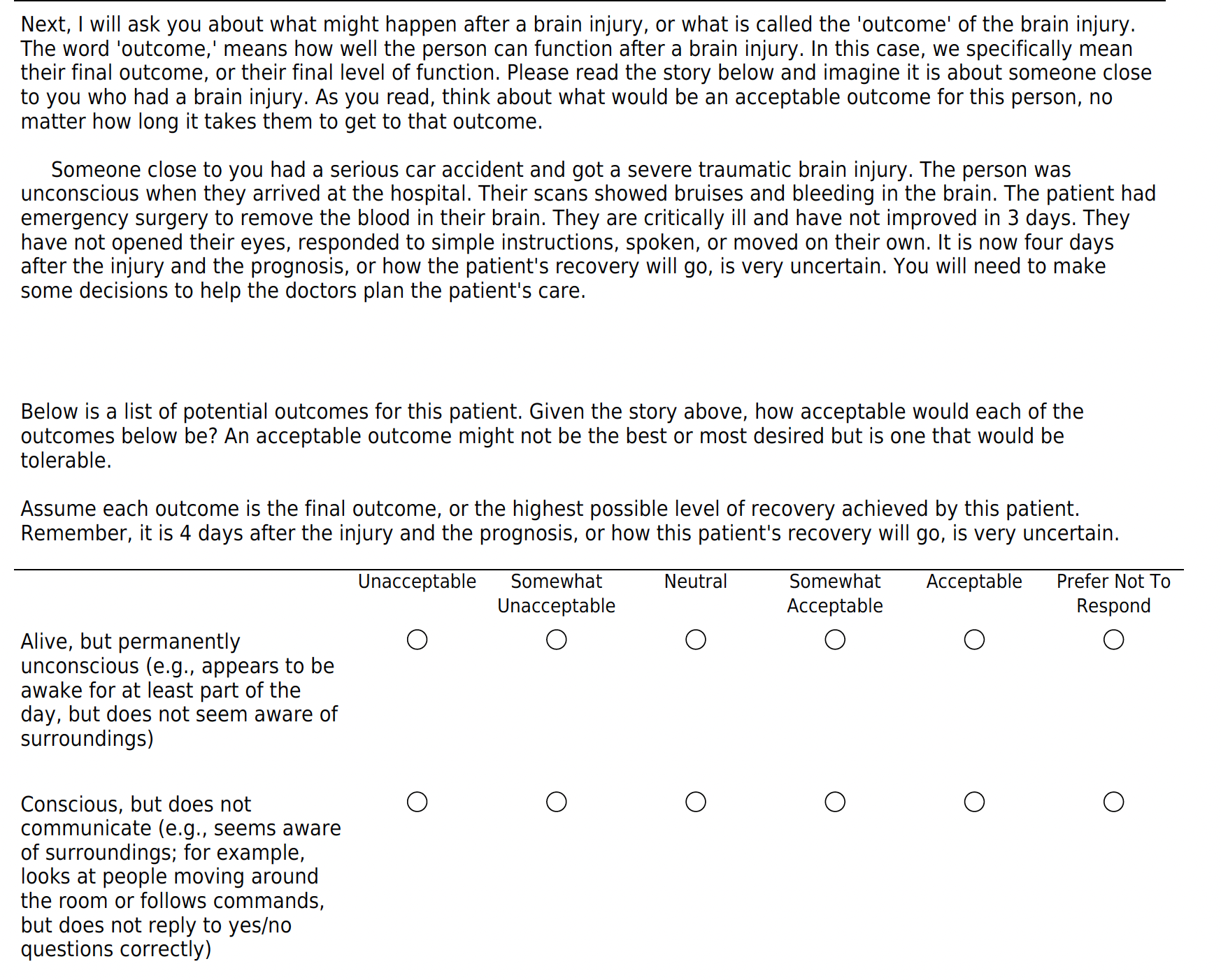

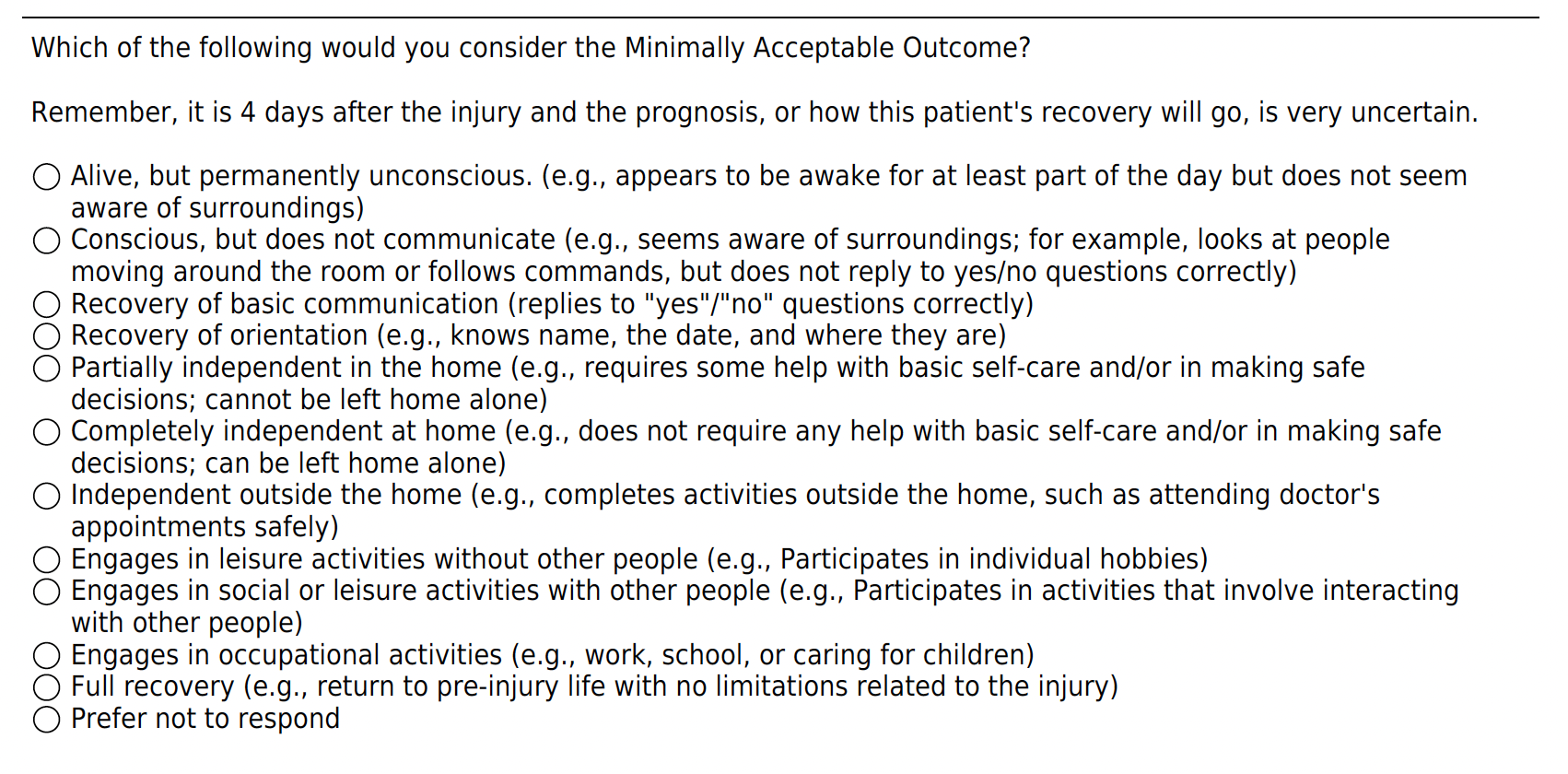

**Supplementary Figure 1: Survey excerpts** Sample questions related to TBI outcome acceptability and minimally acceptable outcome (MAO)

**Supplementary Figure 2**

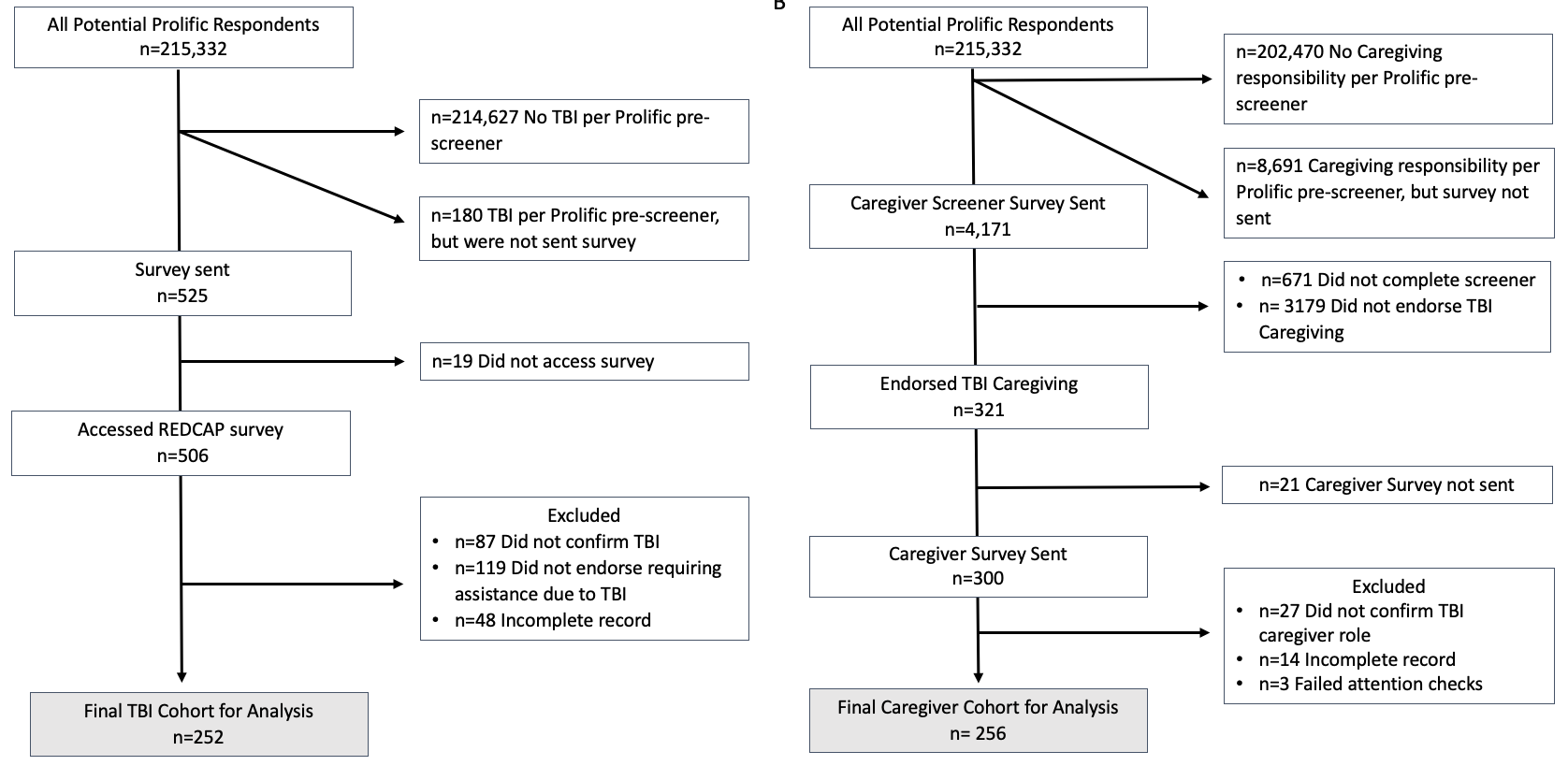

**Supplementary Figure 2: Participant inclusion flow diagrams:** We provided the SPEAC-TBI survey to respondents registered and vetted by Prolific who endorsed having a TBI (A) or a TBI caregiver role (B). To identify persons with TBI registered with the Prolific online platform (A), we used the TBI screener question that is embedded within the Prolific system to exclude persons who did not endorse TBI; 705 Prolific participants endorsed TBI. We sent the SPEAC-TBI survey in batches to a subset of participants endorsing TBI so as not to exceed the enrollment target of 250. We excluded those who did not access the survey, did not confirm having a TBI, did not endorse needing assistance for the TBI, and accessed but did not complete the survey. In total, we sent the survey to 525/705 Prolific participants to obtain our final target of at least 250 complete surveys. We followed a similar procedure to identify TBI caregivers registered with the Prolific online platform (B), however, because the caregiver screener question that is embedded within Prolific is not specific to TBI caregiving, we created a second screener to identify, among Prolific participants who endorsed having a caregiver role, those who provided caregiving to someone with TBI. We released this screener in batches and monitored responses sending the SPEAC-TBI Caregiver survey to those who endorsed TBI caregiving. We excluded participants those who did not complete the screener questions, did not confirm their caregiver role, or failed the attention checks on the SPEAC survey.

**Supplementary Figure 3**
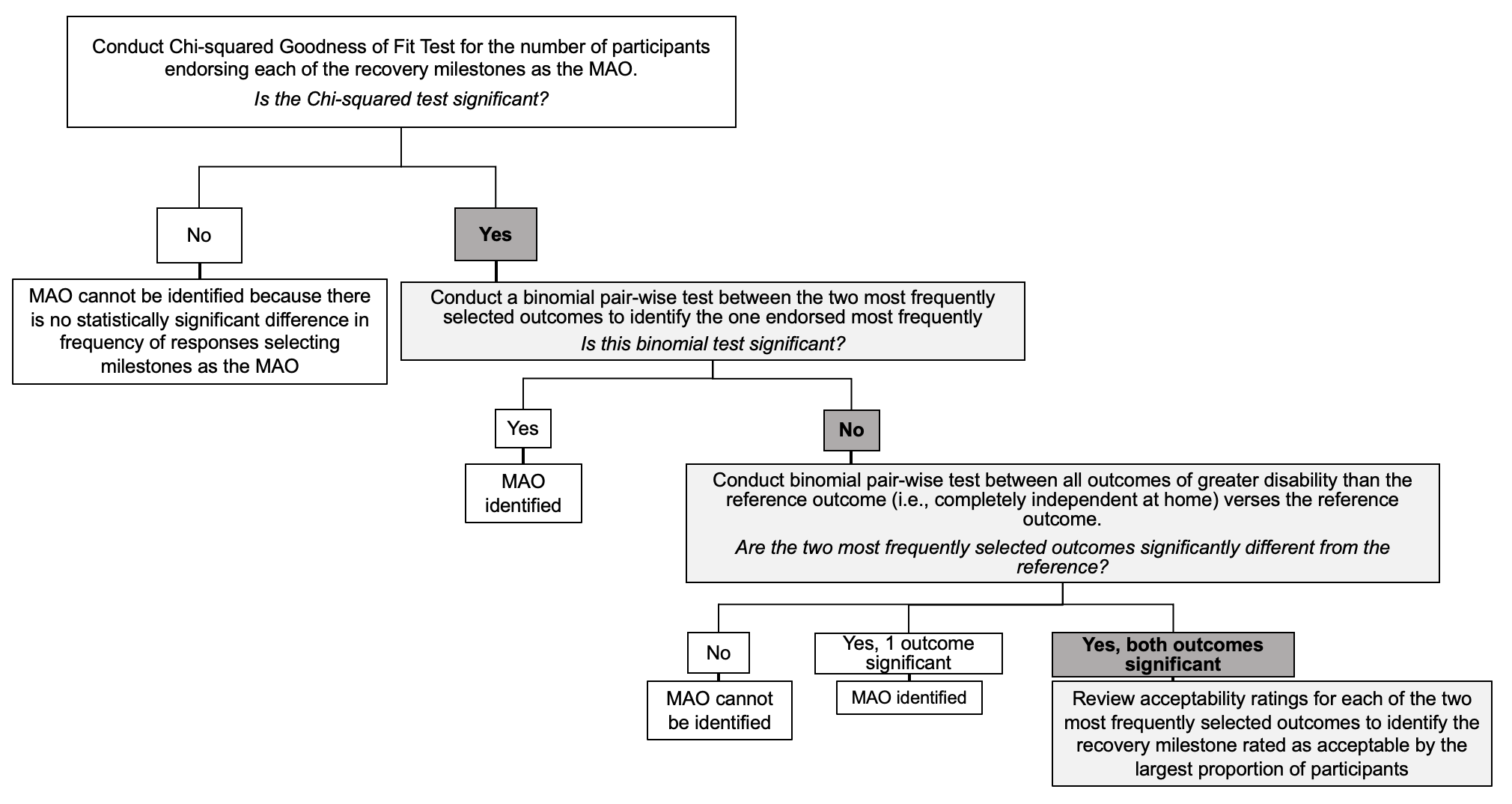

**Supplementary Figure 3: Analytic decision tree** The procedure for selecting the minimally acceptable outcome (MAO) included identifying the two most frequently selected MAO’s by each cohort, determinig whether there was a significant difference in the proportion of participants selecting the most versus the second most selected MAO, and, in the absence of a significnt difference, confirming that both MAO’s were selected more frequently than the reference standard, and using the acceptability ratings to select the final MAO. Grey shading indicates the selection made in each step.

|  | **Supplementary Table 1: Outcome Acceptability Ratings for Persons with TBI and TBI Caregivers** | | | | | | | | | | |
| --- | --- | --- | --- | --- | --- | --- | --- | --- | --- | --- | --- |
|  |  | **Rating** | | | | | | | | | |
|  | **TBI Outcome** | **Unacceptable** | | **Somewhat Unacceptable** | | **Neutral** | | **Somewhat Acceptable** | | **Acceptable** | |
|  |  | **TBI** | **CG** | **TBI** | **CG** | **TBI** | **CG** | **TBI** | **CG** | **TBI** | **CG** |
| **1** | **Alive, permanently unconscious** | 116 (46.2) | 98 (39.2) | 55 (21.9) | 48 (19.2) | 30 (12.0) | 33 (13.2) | 29 (11.6) | 35 (14.0) | 21 (8.4) | 36 (14.4) |
| **2** | **Conscious, does not comm.** | 48 (19.1) | 46 (18.3) | 68 (27.1) | 62 (24.6) | 41 (16.3) | 39 (15.5) | 63 (25.1) | 71 (28.2) | 31 (12.4) | 34 (13.5) |
| **3** | **Recovery of basic comm.** | 7 (2.8) | 9 (3.5) | 49 (19.7) | 25 (9.8) | 30 (12.1) | 38 (14.9) | 73 (29.3) | 82 (32.2) | 90 (36.1) | 101 (39.6) |
| **4** | **Recovery of orientation** | 6 (2.4) | 6 (2.4) | 28 (11.2) | 14 (5.5) | 29 (11.7) | 33 (13.0) | 67 (26.9) | 80 (31.6) | 119 (47.8) | 120 (47.4) |
| **5** | **Partially indep. in the home** | 6 (2.4) | 6 (2.4) | 13 (5.2) | 11 (4.3) | 15 (6.0) | 19 (7.5) | 83 (33.3) | 78 (30.7) | 132 (53.0) | 140 (55.1) |
| **6** | **Completely indep. at home** | 12 (4.9) | 18 (7.1) | 3 (1.2) | 11 (4.3) | 8 (3.3) | 13 (5.1) | 25 (10.2) | 33 (13.0) | 198 (80.5) | 179 (70.5) |
| **7** | **Indep. outside the home** | 11 (4.5) | 15 (6.0) | 5 (2.1) | 13 (5.2) | 8 (3.3) | 12 (4.8) | 12 (4.9) | 39 (15.5) | 207 (85.2) | 172 (68.5) |
| **8** | **Engages in activities without other people** | 7 (2.9) | 11 (4.4) | 8 (3.3) | 15 (6.0) | 5 (2.0) | 11 (4.4) | 14 (5.7) | 31 (12.3) | 211 (86.1) | 184 (73.0) |
| **9** | **Engages in activities with other people** | 5 (2.0) | 8 (3.2) | 6 (2.5) | 8 (3.2) | 9 (3.7) | 19 (7.6) | 14 (5.7) | 25 (10.0) | 210 (86.1) | 191 (76.1) |
| **10** | **Engages in occupational activities** | 9 (3.7) | 14 (5.6) | 7 (2.9) | 12 (4.8) | 11 (4.5) | 10 (4.0) | 14 (5.7) | 32 (12.7) | 204 (83.3) | 184 (73.0) |
| **11** | **Full recovery** | 10 (4.1) | 16 (6.4) | 4 (1.7) | 9 (3.6) | 9 (3.7) | 13 (5.2) | 11 (4.5) | 21 (8.4) | 208 (86.0) | 191 (76.4) |
| Values represent number and proportion of participants n (%); Sample sizes vary slightly due to responses of “Prefer Not to Respond”; CG = caregiver respondents; TBI = respondents with traumatic brain injury | | | | | | | | | | | |

| **Supplementary Table 2: Outcome Acceptability Ratings by Persons with TBI Endorsing Needing Help with Basic Activities Some of the Time Versus All of the Time** | | | |
| --- | --- | --- | --- |
| **TBI Outcome** | **All Persons with TBI**  **N (%)** | **Persons with TBI Who Endorsed Needing Help with Basic Activities *Some of the Time***  **N (%)** | **Persons with TBI Who Endorsed Needing Help with Basic Activities**  ***All of the Time***  **N (%)** |
| **Alive, but permanently unconscious** | | | |
| ***N*** | ***251*** | ***198*** | ***53*** |
| Unacceptable | 116 (46.2) | 89 (44.9) | 27 (50.9) |
| Somewhat Unacceptable | 55 (21.9) | 48 (24.2) | 7 (13.2) |
| Neutral | 30 (12.0) | 27 (13.6) | 3 (5.7) |
| Somewhat Acceptable | 29 (11.6) | 20 (10.1) | 9 (17.0) |
| Acceptable | 21 (8.4) | 14 (7.1) | 7 (13.2) |
| **Conscious, but does not communicate** | | | |
| ***N*** | ***251*** | ***198*** | ***53*** |
| Unacceptable | 48 (19.1) | 40 (20.2) | 8 (15.1) |
| Somewhat Unacceptable | 68 (27.1) | 54 (27.3) | 14 (26.4) |
| Neutral | 41 (16.3) | 33 (16.7) | 8 (15.1) |
| Somewhat Acceptable | 63 (25.1) | 46 (23.2) | 17 (32.1) |
| Acceptable | 31 (12.4) | 25 (12.6) | 6 (11.3) |
| **Recovery of basic communication** | | | |
| ***N*** | ***249*** | ***196*** | ***53*** |
| Unacceptable | 7 (2.8) | 6 (3.1) | 1 (1.9) |
| Somewhat Unacceptable | 49 (19.7) | 39 (19.9) | 10 (18.9) |
| Neutral | 30 (12.0) | 25 (12.8) | 5 (9.4) |
| Somewhat Acceptable | 73 (29.3) | 58 (29.6) | 15 (28.3) |
| Acceptable | 90 (36.1) | 68 (34.7) | 22 (41.5) |
| **Recovery of orientation** | | | |
| ***N*** | ***249*** | ***197*** | ***52*** |
| Unacceptable | 6 (2.4) | 5 (2.5) | 1 (1.9) |
| Somewhat Unacceptable | 28 (11.2) | 21 (10.7) | 7 (13.5) |
| Neutral | 29 (11.6) | 21 (10.7) | 8 (15.4) |
| Somewhat Acceptable | 67 (26.9) | 56 (28.4) | 11 (21.2) |
| Acceptable | 119 (47.8) | 94 (47.7) | 25 (48.1) |
| **Partially independent in the home** | | | |
| ***N*** | ***249*** | ***197*** | ***52*** |
| Unacceptable | 6 (2.4) | 4 (2.0) | 2 (3.8) |
| Somewhat Unacceptable | 13 (5.2) | 11 (5.6) | 2 (3.8) |
| Neutral | 15 (6.0) | 12 (6.1) | 3 (5.8) |
| Somewhat Acceptable | 83 (33.3) | 67 (34.0) | 16 (30.8) |
| Acceptable | 132 (53.0) | 103 (52.3) | 29 (55.8) |
| **Completely independent at home** | | | |
| ***N*** | ***246*** | ***194*** | ***52*** |
| Unacceptable | 12 (4.9) | 9 (4.6) | 3 (5.8) |
| Somewhat Unacceptable | 3 (1.2) | 3 (1.5) | 0 (0.0) |
| Neutral | 8 (3.3) | 6 (3.1) | 2 (3.8) |
| Somewhat Acceptable | 25 (10.2) | 19 (9.8) | 6 (11.5) |
| Acceptable | 198 (80.5) | 157 (80.9) | 41 (78.8) |

| **Supplementary Table 2, con’t** | | | |
| --- | --- | --- | --- |
| **TBI Outcome** | **All Persons with TBI N (%)** | **Persons with TBI Who Endorsed Needing Help with Basic Activities *Some of the Time***  **N (%)** | **Persons with TBI Who Endorsed Needing Help with Basic Activities**  ***All of the Time***  **N (%)** |
| **Independent outside the home** | | | |
| ***N*** | ***243*** | ***192*** | ***51*** |
| Unacceptable | 11 (4.5) | 8 (4.2) | 3 (5.9) |
| Somewhat Unacceptable | 5 (2.1) | 4 (2.1) | 1 (2.0) |
| Neutral | 8 (3.3) | 7 (3.6) | 1 (2.0) |
| Somewhat Acceptable | 12 (4.9) | 9 (4.7) | 3 (5.9) |
| Acceptable | 207 (85.2) | 164 (85.4) | 43 (84.3) |
| **Engages in leisure activities without other people** | | | |
| ***N*** | ***245*** | ***193*** | ***52*** |
| Unacceptable | 7 (2.9) | 4 (2.1) | 3 (5.8) |
| Somewhat Unacceptable | 8 (3.3) | 7 (3.6) | 1 (1.9) |
| Neutral | 5 (2.0) | 5 (2.6) | 0 (0.0) |
| Somewhat Acceptable | 14 (5.7) | 12 (6.2) | 2 (3.8) |
| Acceptable | 211 (86.1) | 165 (85.5) | 46 (88.5) |
| **Engages in social or leisure activities with other people** | | | |
| ***N*** | ***244*** | ***193*** | ***51*** |
| Unacceptable | 5 (2.0) | 3 (1.6) | 2 (3.9) |
| Somewhat Unacceptable | 6 (2.5) | 5 (2.6) | 1 (2.0) |
| Neutral | 9 (3.7) | 9 (4.7) | 0 (0.0) |
| Somewhat Acceptable | 14 (5.7) | 12 (6.2) | 2 (3.9) |
| Acceptable | 210 (86.1) | 164 (85.0) | 46 (90.2) |
| **Engages in occupational activities** | | | |
| ***N*** | ***245*** | ***193*** | ***52*** |
| Unacceptable | 9 (3.7) | 7 (3.6) | 2 (3.8) |
| Somewhat Unacceptable | 7 (2.9) | 4 (2.1) | 3 (5.8) |
| Neutral | 11 (4.5) | 11 (5.7) | 0 (0.0) |
| Somewhat Acceptable | 14 (5.7) | 11 (5.7) | 3 (5.8) |
| Acceptable | 204 (83.3) | 160 (82.9) | 44 (84.6) |
| **Full recovery** | | | |
| ***N*** | ***242*** | ***192*** | ***50*** |
| Unacceptable | 10 (4.1) | 7 (3.6) | 2 (6.0) |
| Somewhat Unacceptable | 4 (1.7) | 4 (2.1) | 0 (0.0) |
| Neutral | 9 (3.7) | 7 (3.6) | 2 (4.0) |
| Somewhat Acceptable | 11 (4.5) | 9 (4.7) | 2 (4.0) |
| Acceptable | 208 (86.0) | 165 (85.9) | 43 (86.0) |
| Sample sizes vary slightly due to selection of the “Prefer not to respond” answer option | | | |

| **Supplementary Table 3: MAOs Selected by Persons with TBI and TBI Caregivers With Low Response Categories Collapsed** | | | |
| --- | --- | --- | --- |
|  | **TBI Outcome** | **Persons**  **with TBI**  **N (%)**  **N = 250** | **TBI Caregiver**  **N (%)**  **N = 255** |
| **1** | Alive, but permanently unconscious | 36 (14.4) | 53 (20.8) |
| **2** | Conscious, but does not communicate | 65 (26.0) | 52 (20.4) |
| **3** | Recovery of basic communication | 54 (21.6) | 57 (22.4) |
| **4** | Recovery of orientation | 27 (10.8) | 27 (10.6) |
| **5** | Partially independent in the home | 46 (18.4) | 42 (16.5) |
| **6** | Completely independent at home | 14 (5.6) | 10 (3.9) |
| **7** | Independent outside the home **through full recovery** | 8 (3.2) | 14 (5.5) |
| Categories: Engages in leisure activities without other people, Engages in social or leisure activities with other people, Engages in occupational activities, and Full recovery were selected by <10 respondents and were collapsed into the “Independent outside the home” category for all statistical analyses. **N=2 (<1%) persons with TBI and N=1 (<1%) TBI caregivers preferred not to respond to this item.** MAO = minimally acceptable outcome | | | |

| **Supplementary Table 4: MAOs Selected by Persons with TBI Endorsing Needing Help with Basic Activities Some of the Time Versus All of the Time** | | | | |
| --- | --- | --- | --- | --- |
|  | **TBI Outcome** | **All Persons**  **with TBI**  ***N = 250***  **N (%)** | **Persons**  **with TBI Who Indicated Needing Help with Basic Activities *Some of the Time***  ***N = 197***  **N (%)** | **Persons**  **with TBI Who Indicated Needing Help with Basic Activities *All of the Time***  ***N = 53***  **N (%)** |
| 1 | Alive, but permanently unconscious | 36 (14.4) | 27 (13.7) | 9 (17.0) |
| 2 | Conscious, but does not communicate | 65 (26.0) | 51 (25.9) | 14 (26.4) |
| 3 | Recovery of basic communication | 54 (21.0) | 42 (21.3) | 12 (22.6) |
| 4 | Recovery of orientation | 27 (10.8) | 20 (10.2) | 7 (13.2) |
| 5 | Partially independent in the home | 46 (18.4) | 38 (19.3) | 8 (15.1) |
| 6 | Completely independent at home | 14 (5.6) | 13 (6.6) | 1 (1.9) |
| 7 | Independent outside the home | 1 (0.4) | 1 (0.5) | 0 (0.0) |
| 8 | Engages in leisure activities without other people | 1 (0.4) | 1 (0.5) | 0 (0.0) |
| 9 | Engages in social or leisure activities with other people | 2 (0.8) | 1 (0.5) | 1 (1.9) |
| 10 | Engages in occupational activities | 0 (0.0) | 0 (0.0) | 0 (0.0) |
| 11 | Full recovery | 4 (1.6) | 3 (1.5) | 1 (1.9) |
| MAO = minimally acceptable outcome  N**=2 (<1%) persons with TBI preferred not to respond to this item** | | | | |

| **Supplementary Table 5: Comparison of the Proportion of Participants Selecting Each Outcome Milestone as the MAO relative to “Completely Independent at Home”** | | | |
| --- | --- | --- | --- |
|  | **TBI Outcome** | **Persons**  **with TBI** | **TBI Caregivers** |
| 1 | Alive, but permanently unconscious | Z = 3.11, p = 0.0019 | Z = 5.42, p < 0.0001 |
| 2 | Conscious, but does not communicate | Z = 5.74, p < 0.0001 | Z = 5.33, p < 0.0001 |
| 3 | Recovery of basic communication | Z = 4.85, p < 0.0001 | Z = 5.74, p < 0.0001 |
| 4 | Recovery of orientation | Z = 2.03, p = 0.0423 | Z = 2.80, p = 0.0038 |
| 5 | Partially independent in the home | Z = 4.13, p < 0.0001 | Z = 4.44, p < 0.0001 |
| 6 | Completely independent at home | Reference | |
| 7 | Independent outside the home **through full recovery** | Z = 1.28, p = 0.2008 | Z = -0.82, p = 0.4142 |
| MAO = minimally acceptable outcome  Reference standard: completely independent at home | | | |
